# Experience of Service Questionnaire (ESQ) in children and adolescents: factor structure, reliability, validity, item parameters and interpretability of the parent version for practical use in Greece

**DOI:** 10.1101/2024.07.05.24309986

**Authors:** Konstantinos Kotsis, Andromachi Mitropoulou, Alexandra Tzotzi, Lauro Estivalete Marchionatti, Mauricio Scopel Hoffmann, Julia Luiza Schafer, Caio B. Casella, André Simioni, Katerina Papanikolaou, Maria Basta, Aspasia Serdari, Anastasia Koumoula, Giovanni Abrahão Salum

## Abstract

**Background:** Health systems need tools to assess patient’s experience of service, but existing tools lack reliability and validity assessment. Our aim is to investigate the factor structure, reliability, validity, item parameters and interpretability of the parent version of the Experience of Service Questionnaire (ESQ) for practical use in Greece.

**Methods:** A total of 265 caregivers that were using mental health services in Greece participated in this study as part of the Nationwide cross-sectional survey from the Child and Adolescent Mental Health Initiative (CAMHI). Confirmatory Factor Analysis was used to test factor structure. Reliability of all models were measured with omega coefficients. Tobit regression analysis was used to test for convergent and discriminant validity with specifically designed questions. Item parameters were assessed via Item Response Theory. Interpretability was assessed by means of IRT-based scores.

**Results:** We found that ESQ is best represented and scored as a unidimensional construct, given potential subscales would not have enough reliability apart from a general factor. Convergent and discriminant validity was demonstrated, as caregivers who perceived that their child benefited from the received mental health care had 6.50 higher summed scores (SMD=1.14, p<0.001); while those who believed that their child needed additional help had 5.08 lower summed scores on the ESQ (SMD=-0.89, p<0.001). Average z-scores provided five meaningful categories of services, in terms of user satisfaction, compared to the national average.

**Conclusions:** Our study presents evidence for the reliability and validity of the ESQ and provides recommendations for its practical use in Greece. ESQ can be used to measure experience of service and might help drive improvements in service delivery in the Greek mental health sector.

## Introduction

Every health care service has the legal and ethical necessity to provide high-quality care. User experiences can be used to assess and benchmark the quality of services or to make institutional improvements [1]. In the case of mental health, better experiences when receiving care have been associated with lower dropout rates and higher engagement in therapy [2–5], leading to positive outcomes through increased involvement [6,7]. Therefore, understanding how users experience mental health services may play a pivotal role in improving quality of care and ensuring patient-centered service delivery.

One essential requirement for improving the quality of care of children and adolescents’s mental health services is reliable measurement through valid and informative tools. Patient-reported experience measures assess a healthcare provider’s services from the patients’ or their proxies’ viewpoints [8]. There are a few studies [1,9–11] exploring psychometric properties of various tools measuring satisfaction in Child and Adolescent Mental Health Services (CAMHS) such as Broad Evaluation of Satisfaction with Treatment (BEST), CAMHS Satisfaction Scale (CAMHSSS), Parent Experiences Questionnaire for Outpatient CAMHS (PEQ-CAMHS), and Experience of Service Questionnaire (ESQ). ESQ represents the core measure for service experience across many CAMHS in the UK, it has been developed by the Commission for Health Improvement considering users’ experience (parents and children) in well-designed focus groups, and it is a brief questionnaire (12 items) and free to use.

To the best of our knowledge, only the original study [1] provides validation data for the ESQ, while other studies [12–14] explore satisfaction levels, association with clinical outcomes, and predictors. According to the authors of the original study, two related constructs were measured by the ESQ: satisfaction with care and satisfaction with environment. Authors suggest that these two constructs represent related aspects of global satisfaction, supporting the view that responses to patient-reported experience measures are universally represented by a general attribute of satisfaction.

The question on whether ESQ measures one or two related constructs is an important one for the practical use of the tool. Model fit has been criticized as the sole parameter for deciding on scoring a measurement tool [15,16]. Several times, even when tools can have a better fit by adding more dimensions, this does not mean that dimensions have enough indicators for reliably scoring practices. One way to address this issue is by investigating reliability with bifactor models. Bifactor models separate general from specific variance and are able to investigate if after accounting for a general factor, whether there is enough variability for scoring subscale scores. Given the results just outlined by the original validated study, it is important to further explore if ESQ has enough variability in subscores to allow rating in two scales; or whether ESQ is better scored as a single score. In the latter case, care and environment seem to measure separate aspects but they are so related that different scoring would not be sufficiently reliable.

In the context of Greece, there is limited availability of feedback tools to measure user experience in mental health services. Due to the differences in mental health systems among countries and the fact that the concept of “satisfaction” might be influenced by social, financial and cultural factors [17], it is important to explore user’s experience at a country level with context-sensitive tools. However, there are no tools specifically tailored for children and adolescents, and international patient-reported experience measures are not translated and/or validated for the Greek population, creating barriers for consistent and reliable assessment of service quality.

The aim of our study is to confirm the factor structure, the reliability (internal consistency), construct validity (convergent and discriminant) and interpretability of the ESQ in a nationwide sample of caregivers whose children are receiving mental health care in Greece. We hypothesize that ESQ will be a reliable tool that measures satisfaction that should be scored as a single domain. We extend prior work by exploring its construct convergent and discriminant validity and use item response theory to aid the interpretability of ESQ in Greece. We expect that a scoring system which can be used to evaluate CAMHS, by considering parental feedback, will consequently help improving services.

## Methods

### Participants

We used data from a 2022/2023 cross-sectional survey from the Child and Adolescent Mental Health Initiative (CAMHI) on the current state and needs for child and adolescent mental health in Greece based on multiple viewpoints [18]. A nationwide sample of 1,756 caregivers participated in the online survey, answering questions related to service use and access, literacy and stigma, parenting practices, and mental health needs of their children/adolescents. Out of them, 295 caregivers who had visited a Child and Adolescent Mental Health Service answered the ESQ-parent version. Recruitment occurred through an online respondent panel provided by the research company IQVIA OneKey, which was developed based on census quotas, reaching participants online via social media and website campaigns, search engine optimization, panelists’ friends referrals, and affiliate networks [19]. To avoid self-selection, the online surveys were automatically routed to respondents based on a specific algorithm. Data was collected and preserved according to the General Data Protection Regulation (GDPR) National Policy [20]. Ethical approval was granted by the Research Ethics Committee of the Democritus University of Thrace [approval number: ΔΠΘ/EHΔE/42772/307].

### Instrument

#### Experience of Service Questionnaire (ESQ) - parent version

the instrument was developed to assess parents’ and children’s positive experiences from mental health services (in this study, we solely focused on parents’ experience). The original study showed ESQ is best captured as two related constructs: Satisfaction with Care (items 1,2,3,4,5,6,7,11,12) and Satisfaction with Environment (items: 8,9,10) [1]. But authors also advise to score only the Satisfaction with Care subscore, given the degree of relatedness between the two constructs. The instrument is rated on a 4-point Likert-type scale (certainly true, partially true, not true, and a last option of “I don’t know”, which was considered a missing variable). Most ESQ data is now collected and stored in a format where 1 = Not true, 2 = Partly true, 3 = Certainly true [21]. Total scores range from 12 to 36 (9 to 27 in the Care subscale and 3 to 9 in the Environment scale), with higher scores representing better service experience as all questions are written as positive statements (e.g. “I feel the people here know how to help with the problem I came for”). There are also three free-text sections looking at what the respondent liked about the service, what they felt needed improving, and any other comments. The ESQ was translated and culturally adapted to Greek following a validated five-stage procedure and is freely available to use [22].

### Statistical analysis

First, we performed Confirmatory Factor Analysis (CFA) to evaluate the factor structure of ESQ based on the correlated model (Satisfaction with care and Satisfaction with environment) described in the original study [1]. Given the original study suggested very high correlation scores between the two constructs, we also explored model fit for three alternative models: the unidimensional model (all items loading into a general factor), a second order model (satisfaction with care and satisfaction with environment as lower order factors and overall satisfaction as a high order factor) and a bifactor model with one general satisfaction factor and two specific factors (Specific satisfaction with care and Specific satisfaction with environment). Global model fit was evaluated with the following fit indices: the Comparative Fit Index (CFI), the Tucker-Lewis Index (TLI), the Root Mean Square Error of Approximation (RMSEA), and the Standardized Root Mean-square Residual (SRMR). A good fit is indicated by the following values: SRMR < 0.6; RMSEA < 0.06; TLI and CFI > 0.95 [23].

Second, our reliability analysis was tested by means of internal consistency (the degree of interrelatedness among the items) [24]. Reliability analysis by area of latent trait was performed using Cronbach alpha and by Omega (ω) coefficient for each ESQ model tested. Cronbach alpha, assumes equal loadings (essential tau equivalence) and a value of 0.7 is considered acceptable [25]. Omega estimates the proportion of variance in the observed total score attributable to all “modeled” sources of common variance. A value of >0.65 for omega total (ω_t_) is considered acceptable and >0.8 is considered strong [26]. For the bifactor model we also assessed omega hierarchical (ω_h_) and omega hierarchical subscale (ω_hs_). Coefficient ω_h_ estimates the proportion of variance in total scores that can be attributed to a single general factor. ω_hs_ is an index reflecting the reliability of a subscale score after controlling for the variance due to the general factor. For ω_h_ values of >0.80 are recommended [26]. To further assess reliability of the factors, we estimate factor determinacy (FD), explained common variance (ECV) and percentage of uncontaminated correlations (PUC). FD estimates the reliability of factor scores from the correlation between a factor and the scores generated from that factor; ECV is the proportion of the total variance in all items explained by the general factor rather than the specific factors and PUC is the percent of all correlations among symptoms attributable purely to the general factor. When ω_H_ is > 0.8 and ECV and PUC are > 0.7, the construct can be interpreted as unidimensional [27]. Higher ECV values indicate a strong general factor, which may guide in the decision to fit a unidimensional model even to data that has evidence of multidimensionality [28].

Third, we tested convergent and discriminant validity, i.e., the degree to which ESQ is consistent with our hypothesis [24]. For that, we performed a Tobit regression analysis, since ESQ total score was right-censored in our data. We investigated the associations between total ESQ score with two variables created by our team as part of the nationwide survey questionnaire: (a) “Do you believe the assistance the child/adolescent has received has helped him/her?” and (b) “Do you believe the child/adolescent needed a different kind of assistance?”. Caregivers had the option to answer these questions if they had answered positively to the question “Has this child/adolescent ever needed any kind of mental health assistance?”. We assumed that the positive answer to question (a) represents a child that has benefitted from the help they had received (convergent validity with ESQ score) and to question (b) represents a child that has not benefited (discriminant validity with ESQ score). We expected that the ESQ score would be associated positively with question (a) and negatively with question (b) given that higher the ESQ score, the better the satisfaction with the service.

Fourth, for interpretability, the degree to which one can assign qualitative meaning to an instrument’s quantitative scores or change in scores [24]ESQ has polytomous response options and therefore the graded response model (GRM) was used to estimate item parameters. We also assess unidimensional item response theory assessments on where ESQ provides information according to the latent trait. Moreover, we estimated the IRT factor scores of the latent variable to rank them into percentiles aiming to provide a meaningful scoring to stakeholders and researchers. To obtain the summed scores of the ESQ constructs we imputed missing values (participants that answered “I don’t know”) with the median score.

Analysis was performed using the software RStudio version 2023.12.1 [29] and the packages *lavaan* [30], *psych* [31], *ltm* [32], *and semTools* [33]. The terms used in this study are following the COSMIN (Consensus-based Standards for the Selection of Health Measurement Instruments) taxonomy of Measurement Properties [24]. Database sheets and the code is openly available at our repository (https://osf.io/crz6h/).

## Results

### Participants

Sample characteristics are shown in Table 1. The majority of the respondents were female (62.0%), and were in a relationship (79.0%). Nearly all participants (97.9%) have finished the mandatory (9 years) education in Greece.

**Table 1.** Sample characteristics (n=295)

|  | Mean | SD |
| --- | --- | --- |
| Caregivers' age | 42.23 | 7.51 |
| Child's age | 11.69 | 4.07 |
|  | **n** | **%** |
| Gender (Female) | 183 | 62.0 |
| Relationship status |  |  |
| Single | 15 | 5.1 |
| Relationship/Cohabitation/Married | 233 | 79.0 |
| Separated/Divorced/Widowed | 47 | 15.9 |
| Educational Level |  |  |
| Mandatory (Grade 1-9) | 6 | 2.1 |
| Non-Mandatory (Grade 10-12) | 110 | 37.3 |
| Higher (Tertiary, MSc, PhD) | 178 | 60.3 |
| Other | 1 | 0.3 |
| Income |  |  |
| Less than 1000€ monthly | 93 | 31.5 |
| Between 1001 to 2,000€ monthly | 108 | 36.6 |
| Above 2000€ monthly | 81 | 27.5 |
| I don't know/ Not applicable | 13 | 4.41 |

### Descriptive statistics

Mean scores of each item are presented in Table 2. All scores are close to 3 (“certainly true”), representing a high satisfaction. Moreover, all items are negatively skewed. The item with the highest score (highest satisfaction) is “I was treated well by the people who have seen my child” and the item with the smallest score (worst satisfaction) is “The appointments are usually at a convenient time (e.g. don’t interfere with work, school)”. A correlation matrix, the histograms of factor score and summed score for the total score as well as the correlation plot of factor scores against summed score are provided in Figure S1, supplementary material.

**Table 2.**
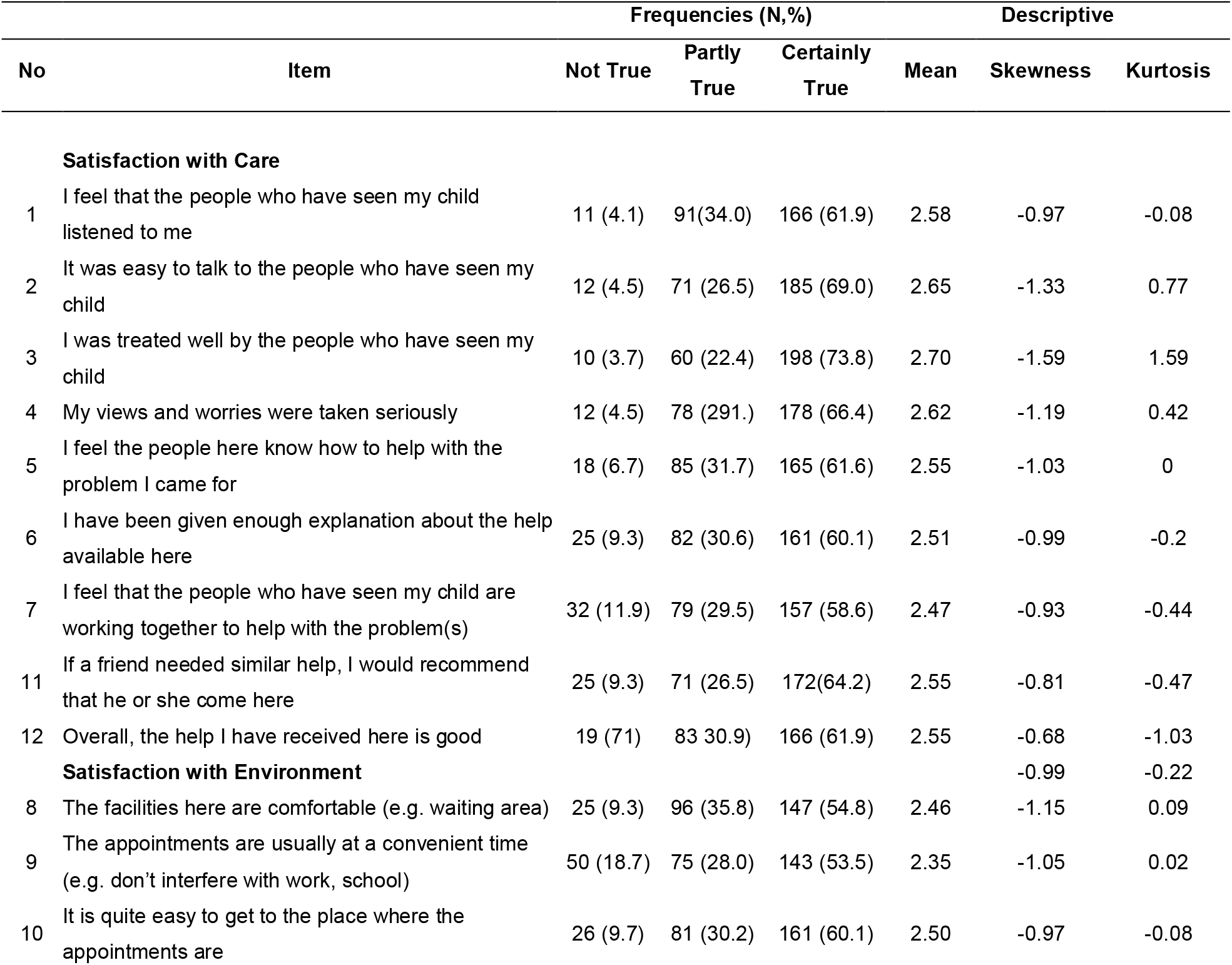
Item descriptive statistics.

|  |  | Frequencies (N,%) |  |  | Descriptive |  |  |
| --- | --- | --- | --- | --- | --- | --- | --- |
| No | Item | Not True | Partly True | Certainly True | Mean | Skewness | Kurtosis |
| Satisfaction with Care |  |  |  |  |  |  |  |
| 1 | I feel that the people who have seen my child listened to me | 11 (4.1) | 91(34.0) | 166 (61.9) | 2.58 | -0.97 | -0.08 |
| 2 | It was easy to talk to the people who have seen my child | 12 (4.5) | 71 (26.5) | 185 (69.0) | 2.65 | -1.33 | 0.77 |
| 3 | I was treated well by the people who have seen my child | 10 (3.7) | 60 (22.4) | 198 (73.8) | 2.70 | -1.59 | 1.59 |
| 4 | My views and worries were taken seriously | 12 (4.5) | 78 (291.) | 178 (66.4) | 2.62 | -1.19 | 0.42 |
| 5 | I feel the people here know how to help with the problem I came for | 18 (6.7) | 85 (31.7) | 165 (61.6) | 2.55 | -1.03 | 0 |
| 6 | I have been given enough explanation about the help available here | 25 (9.3) | 82 (30.6) | 161 (60.1) | 2.51 | -0.99 | -0.2 |
| 7 | I feel that the people who have seen my child are working together to help with the problem(s) | 32 (11.9) | 79 (29.5) | 157 (58.6) | 2.47 | -0.93 | -0.44 |
| 11 | If a friend needed similar help, I would recommend that he or she come here | 25 (9.3) | 71 (26.5) | 172(64.2) | 2.55 | -0.81 | -0.47 |
| 12 | Overall, the help I have received here is good | 19 (71) | 83 30.9) | 166 (61.9) | 2.55 | -0.68 | -1.03 |
| Satisfaction with Environment |  |  |  |  |  | -0.99 | -0.22 |
| 8 | The facilities here are comfortable (e.g. waiting area) | 25 (9.3) | 96 (35.8) | 147 (54.8) | 2.46 | -1.15 | 0.09 |
| 9 | The appointments are usually at a convenient time (e.g. don't interfere with work, school) | 50 (18.7) | 75 (28.0) | 143 (53.5) | 2.35 | -1.05 | 0.02 |
| 10 | It is quite easy to get to the place where the appointments are | 26 (9.7) | 81 (30.2) | 161 (60.1) | 2.50 | -0.97 | -0.08 |

### Factor Structure

The correlated model (*satisfaction with care* and *satisfaction with environment*) showed excellent fit indices to the data (RMSEA = 0.025, CFI = 0.999, TLI = 0.999, SRMR = 0.047) in accordance to the original theoretical construct [1]. Factor loadings were very high, ranging from 0.79 to 0.91 in satisfaction with care, and from 0.78 to 0.89 in satisfaction with environment (Table 3). A high correlation (0.75) was found between the two constructs.

**Table 3.**
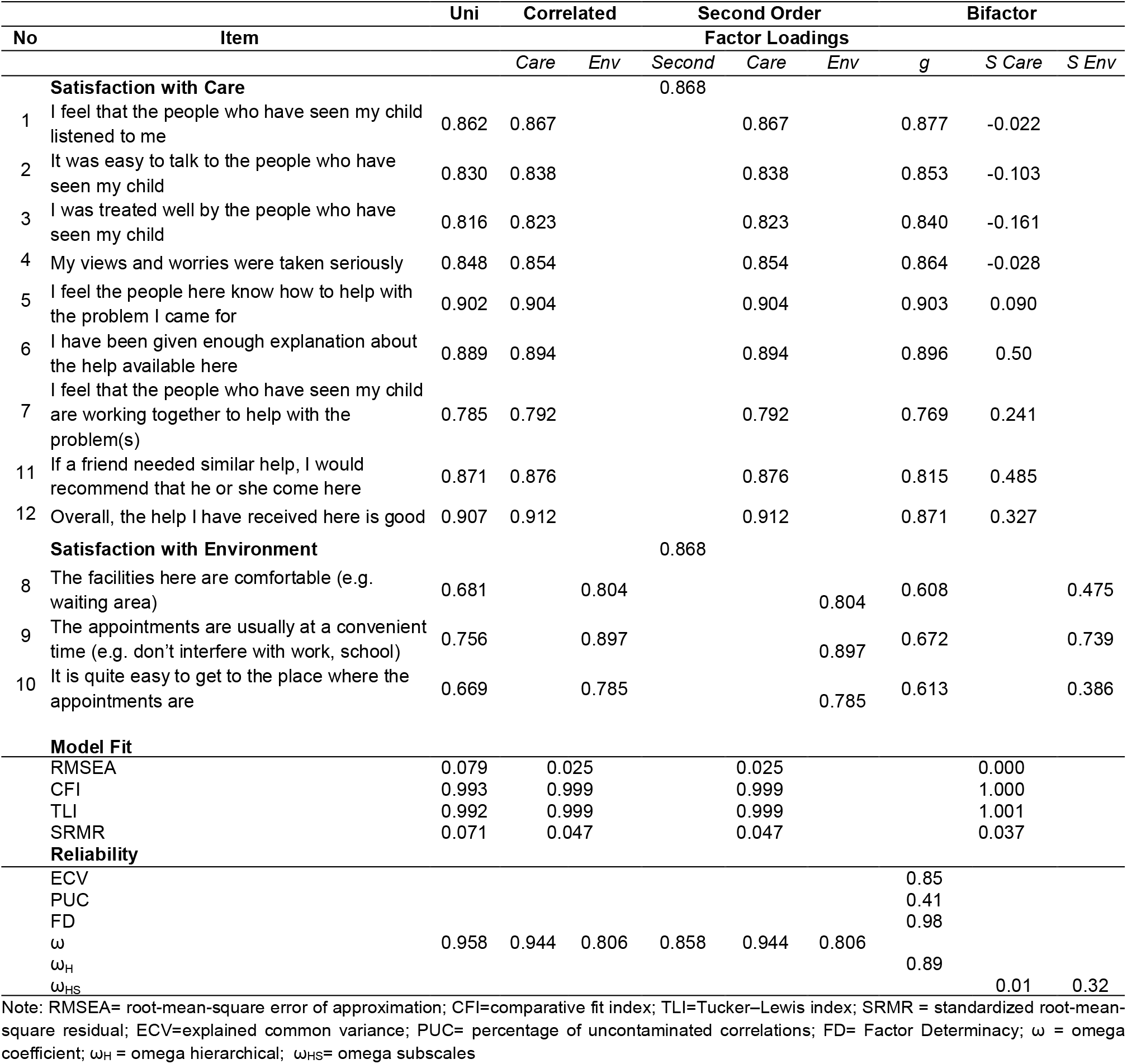
Confirmatory Factor Analysis parameters and reliability coefficients.

Given the high correlation shown in the original paper (and confirmed in the present work), we also examined unidimensional, second order and bifactor models. The unidimensional model revealed that a single factor does capture adequately variance in ESQ scores with acceptable RMSEA value (RMSEA = 0.079) (Table 3). The second order model fitted the data well, suggesting that whereas there are two sources of variance, those two sources can be subsumed under a general overall satisfaction factor, considering the high omega value (Table 3). Finally, the bifactor model revealed that all items load significantly into a strong (given the ECV value) general factor with high factor loadings ranging from 0.61 to 0.90. However, specific factors revealed low and negative factor loadings, which suggests that after accounting for the general factor, the interpretability of specific factors might be compromised, suggesting unidimensionality.

### Reliability

All models presented high reliability with ω coefficients above cut-offs (Table 3). However, the omega hierarchical for the bifactor model was found high (ω_h_=0.89) for the general factor, while omega subscale values for the specific factors were very poor. This suggests that the majority of reliable variance in subscale scores was attributable to the general factor, which precludes meaningful interpretation of subscale scores as unambiguous indicators of a specific factor. Internal consistency by the area of latent trait showed that ESQ is reliable for latent scores ranging from the mean to three standard deviations below the mean. Cronbach’s alpha values were 0.83, 0.87, 0.84 and 0.85 for the mean, one, two and three standard deviations below the mean, respectively. Contrary, reliability was poor for scores above the mean, indicating that the questionnaire is better at capturing information related to poor services in terms of user satisfaction.

### Construct validity

Convergent and discriminant validity was demonstrated for the total satisfaction score (Figure 1). Those who benefited from CAMHS services in Greece had 6.50 higher summed scores (SMD=1.14; t-value = 7.43, p<0.001); while those who believed that their child needed additional help had 5.08 lower summed scores on the same scale (SMD=-0.89, t-value = −7.51, p<0.001). As consistent with a construct validity assessment, this represents high effect sizes.

**Figure 1.**
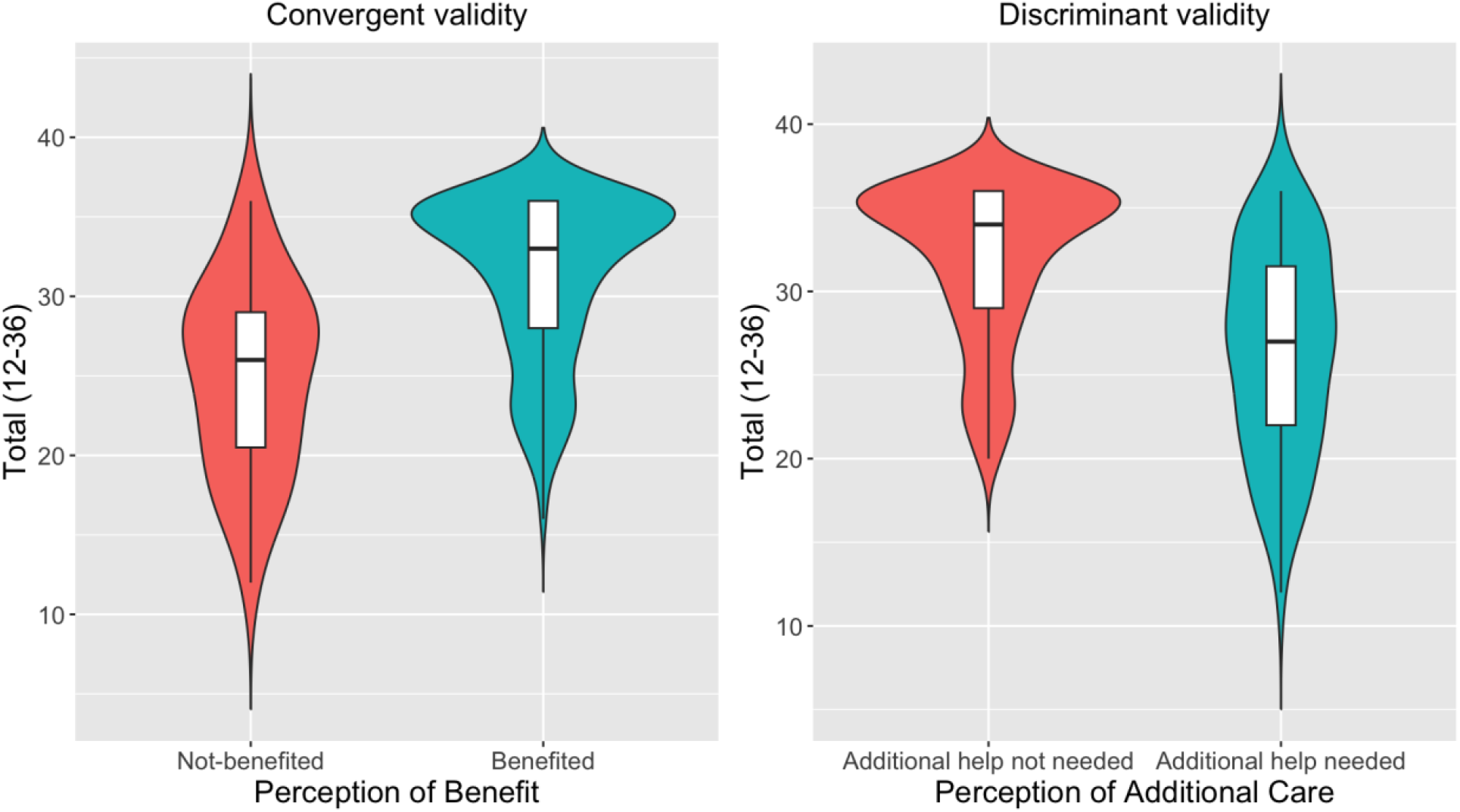
Convergent and discriminant validity comparing ESQ Total Score with perception of benefit and additional care

### Interpretability

#### Unidimensional Item Performance Analysis

Item response function curves and item information curves for each item can be found in Figures S2 and S3, supplemental material. Test information function plot (Figure 2) shows that ESQ provides the most information about slightly-lower-than-average satisfaction levels (the peak is around θ=-0.3) and about slightly-higher-than-two standard deviations below the mean satisfaction levels.

**Figure 2.**
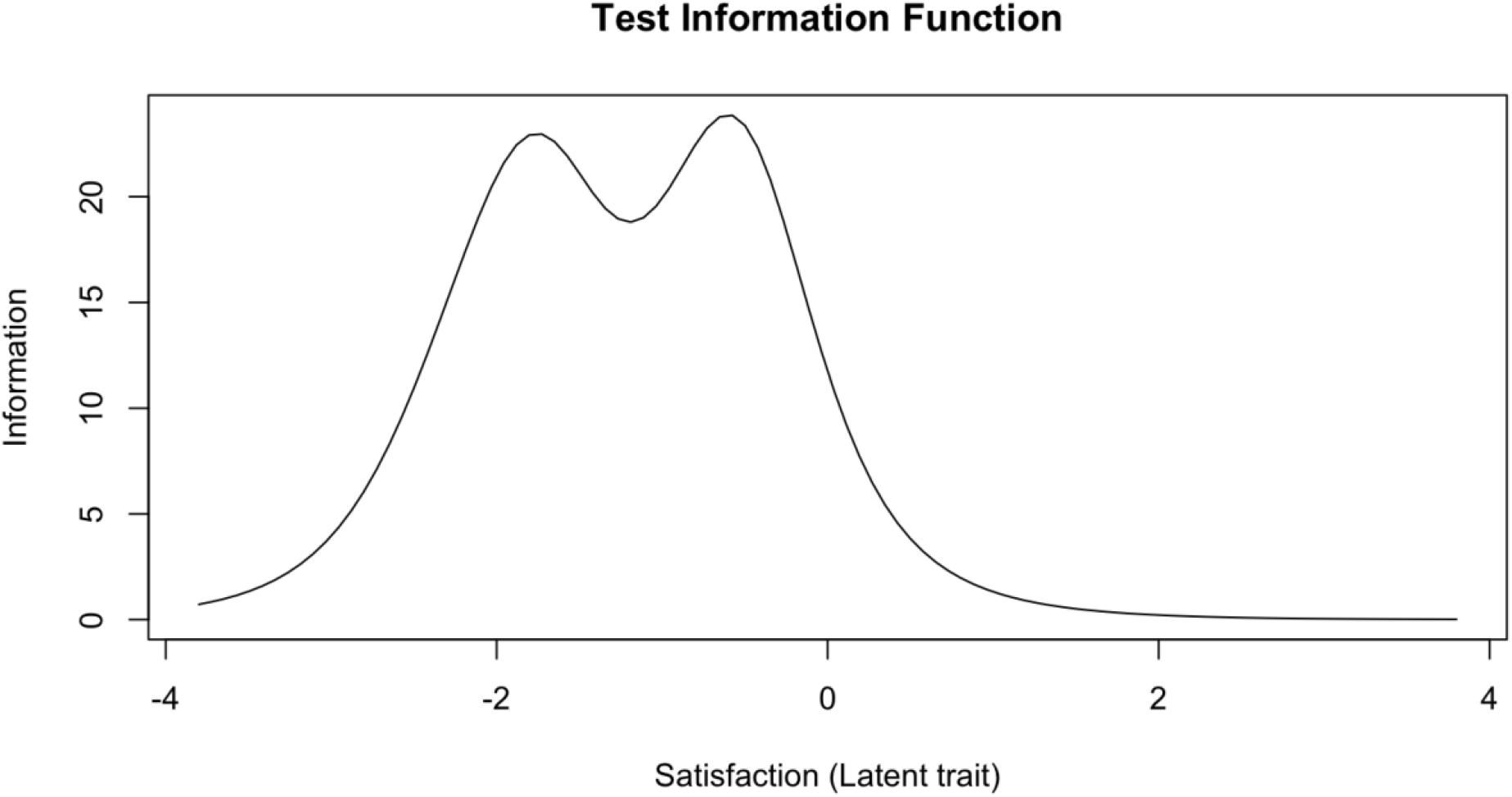
Test Information Function of the Experience of Service Questionnaire (unidimensional solution)

#### Linking summed scores to IRT-based z-scores

The z-scores for the latent variable (Table 4) provide a reference point to assess interpretability of the ESQ. Based on those scores, we classified services as: (1) above average (ESQ total = 36); (2) about average (ESQ total 31-35); (3) slightly below average (ESQ total 26-30); (4) Markedly below average (ESQ total 18-25); and (5) Critically below average (ESQ total 12-17).

**Table 4:**
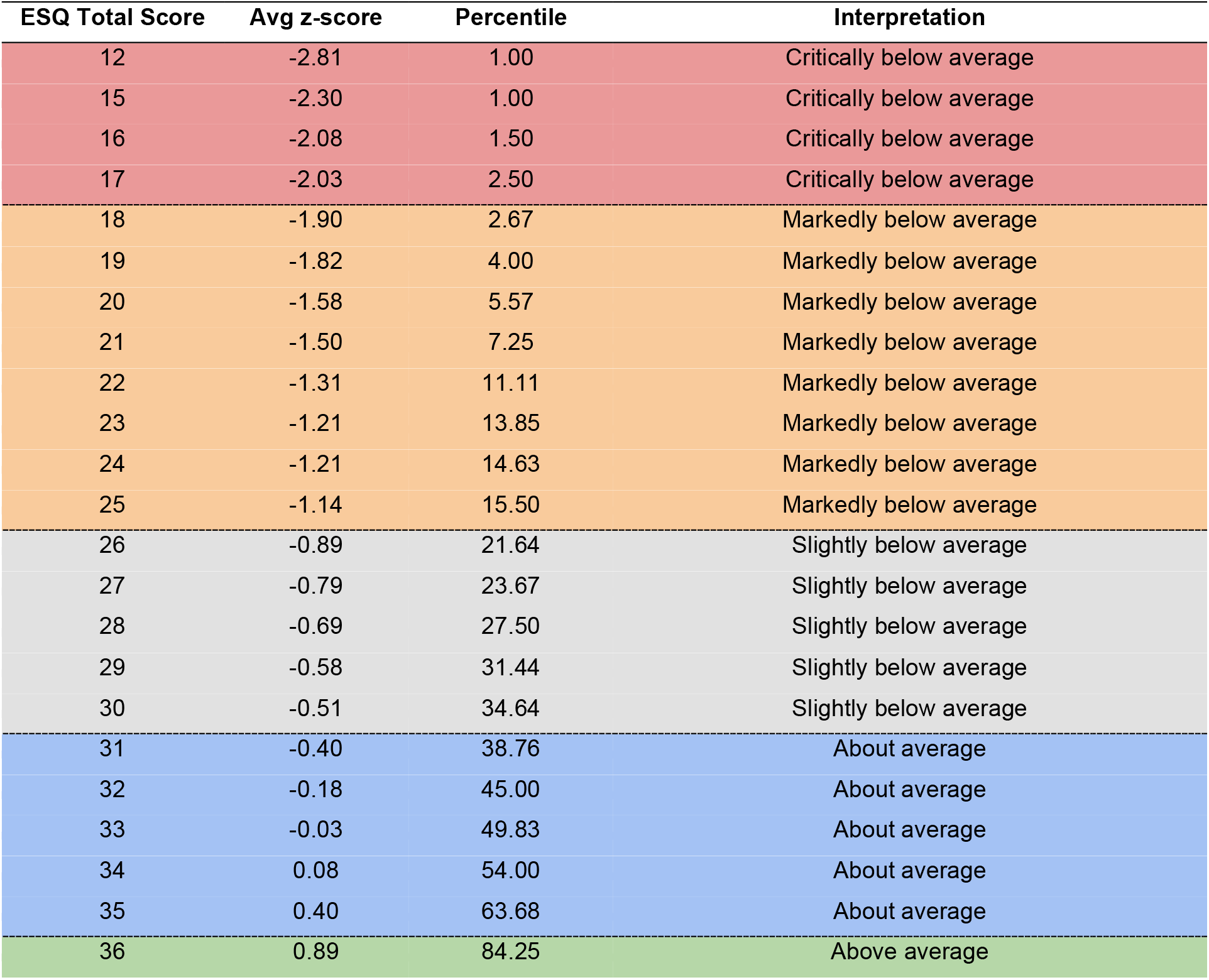
Interpretation of the Experience of Service Questionnaire total score.

| ESQ Total Score | Avg z-score | Percentile | Interpretation |
| --- | --- | --- | --- |
| 12 | -2.81 | 1.00 | Critically below average |
| 15 | -2.30 | 1.00 | Critically below average |
| 16 | -2.08 | 1.50 | Critically below average |
| 17 | -2.03 | 2.50 | Critically below average |
| 18 | -1.90 | 2.67 | Markedly below average |
| 19 | -1.82 | 4.00 | Markedly below average |
| 20 | -1.58 | 5.57 | Markedly below average |
| 21 | -1.50 | 7.25 | Markedly below average |
| 22 | -1.31 | 11.11 | Markedly below average |
| 23 | -1.21 | 13.85 | Markedly below average |
| 24 | -1.21 | 14.63 | Markedly below average |
| 25 | -1.14 | 15.50 | Markedly below average |
| 26 | -0.89 | 21.64 | Slightly below average |
| 27 | -0.79 | 23.67 | Slightly below average |
| 28 | -0.69 | 27.50 | Slightly below average |
| 29 | -0.58 | 31.44 | Slightly below average |
| 30 | -0.51 | 34.64 | Slightly below average |
| 31 | -0.40 | 38.76 | About average |
| 32 | -0.18 | 45.00 | About average |
| 33 | -0.03 | 49.83 | About average |
| 34 | 0.08 | 54.00 | About average |
| 35 | 0.40 | 63.68 | About average |
| 36 | 0.89 | 84.25 | Above average |

## Discussion

The aim of our study was to investigate the factor structure, the reliability (internal consistency), validity and interpretability of the ESQ (parent version) scores in a nationwide sample of caregivers with experiences of mental health services for their children/adolescents in Greece. In accordance with our hypothesis, ESQ proved a reliable tool that measures satisfaction as two strongly related constructs: care and environment. However, a closer inspection of the relationship between the two constructs suggests that after accounting for the general factor, the variance left for specific aspects of care and environment is unreliable, indicating the ESQ is best scored as a single construct. We extend prior work, by demonstrating ESQ has both convergent and discriminant validity and provide practical rules for interpretability by means of using IRT-based scores (z-scores).

The psychometric testing of the ESQ provided good evidence for data quality and internal consistency. The correlated model with the two underlying constructs, as the original study suggests, fits well the data indicating that ESQ can measure parental satisfaction for both care and environment components in a mental health service. However, testing for unidimensionality, and the results derived from unidimensional, second-order and bifactor models indicate that the ESQ item structure is consistent with a single construct. Therefore, we can conclude that the ESQ can measure the satisfaction of parents in Greek CAMHS and we argue for its use, by calculating a single score, rather than subscores for care and environment.

To the best of our knowledge there are no studies exploring the ESQ factor structure in languages other than English. Our results seem to align with the original study [1] in which the authors suggest that responses to service feedback questionnaires are underlain by a common factor, which they interpreted as satisfaction. It is noteworthy that in the original study, the authors found that environment items strongly correlated to each other as well as substantially correlated with the care items. They suggested that there is a strong “halo” effect [1]. Halo effect or affective overtones, is the overgeneralization of characteristics based on one significant dimension [34]. Literature suggests that this occurs when an overall evaluation affects the ratings and therefore underlying perceptions remain covered. This seems true for services, given the fact that when patients share their experience, they do that in either positive or negative way [35]. This observation appears consistent with our study, given the high satisfaction levels reported. Patient-reported experience measures are commonly associated with positive feedback. [35]. However, it’s essential to acknowledge potential biases in sampling and the influence of social desirability effects in satisfaction evaluation studies [36,37]. Concerning the care and environment constructs, authors suggest that they represent related aspects of patients’ satisfaction, and that ESQ should be used as a subjective measure of satisfaction rather than an objective report of care quality or quality of the environment of the service.

Reliability by the area of latent trait was also very high but only for scores below the mean. Therefore, ESQ seems to be able to measure dissatisfaction better than satisfaction indicating that the scale is more reliable when used to identify services that might need improvement. Nonetheless, ESQ represents a reliable measure in the literature. A Norwegian study [13] reported Cronbach’s alpha values of 0.92, 0.93, and 0.61 for general satisfaction, satisfaction with care and satisfaction with the environment, respectively. Additionally, the Spanish version [12] used in parents from Argentina showed an acceptable reliability of α=0.68.

The convergent and discriminant validity was both demonstrated significantly with the perception of parents regarding if the care was beneficial for their child and with the perception of additional help needed. To the best of our knowledge this is the first study demonstrating good concurrent and discriminant external validity of the ESQ, which was a limitation noted in the original study [1]. However, future studies can also benefit from investigating concurrent and discriminant external validity with tools measuring satisfaction in an objective manner, yet not such tools are available in Greece.

Moreover, item performance analysis showed that ESQ is better for parents who are not satisfied at all with the service. Furthermore, items capture more information for respondents with high levels of dissatisfaction. The above findings converge that ESQ is better to capture the lower levels of satisfaction. We could argue, from a service point of view, that this a desirable feature of a patient reported experience measure, since the focus should be in the improvement of not-well running services. Williams et al. [35] pointed out *“that dissatisfaction rather than satisfaction scores may be more useful as an indication of a minimum level of negative experience and in benchmarking exercises”*.

### Limitations and strengths

Our study has important limitations. First of all we did not account for the type of service or the professional in which or by whom the child received mental health care. e.g. public or private, mental health service or mental healthcare in the school setting, psychologist or child and adolescent psychiatrist. This is important since each type of service or professional presents advantages and disadvantages. For example, while most public services in Greece only work till afternoon (thus possibly conflicting with school and parental working hours) and have long waiting lists, they are mainly free and offer multidisciplinary treatment. On the other hand, private practice does not have so long waiting lists, but cost can be an issue, especially when a series of appointments are needed. A second limitation lies in the absence of a valid tool for assessing concurrent and discriminant validation in Greece, yet we are not aware of any other validation of ESQ in the international literature. Finally, we did not account for children’s views to provide an overall validation of ESQ and to explore agreement. To our point of view future studies should incorporate youth’s perspective as well as different types of service (e.g. health - school settings, outpatient - inpatient, well-staffed - understaffed, public - private etc).

Our study also has several strengths that should be emphasized. It represents the first study in the literature providing support about ESQ concurrent and discriminant validity adding, new information to the ESQ literature by providing evidence that the ESQ accurately, indeed, targets satisfaction. Second, item performance analyses provided psychometric evidence of ESQ adequacy on an item-based approach, surpassing the limitations of classical test theory analyses. Moreover, to the best of our knowledge it is the first study measuring satisfaction of parents in Greece concerning CAMHS. The data could be used, in light of the above limitations, for understanding advantages and disadvantages of Greek services and may be used for a baseline information for future studies. Finally, our study provides to the Greek service providers and stakeholders a valid tool to explore client satisfaction and improve their services, if needed. This aligns with the call for measurement in quality of care highlighted by various international stakeholders [38].

## Conclusions

Measuring parental satisfaction is essential for understanding their opinion about received care. The present study supports the use of ESQ in Greek mental health services. The ESQ is valid to measure the general satisfaction of parents by summing the total score. We argue that ESQ can better capture parental dissatisfaction, and that it is a useful measure for service providers in order to improve their care. We acknowledge, however, that satisfaction can vary based on various factors, including individual and family circumstances, as well as contextual factors within the services such as limited staffing and underfunding. Our results suggest that stakeholders can use this information to identify aspects of their services that parents may find dissatisfactory and work toward improvement. Moreover, using this tool at a National level may represent a step forward for Greek services as monitoring satisfaction is lacking in Greece and health policy highlights the importance of capturing clients feedback as a key indicator for the quality of healthcare [39,40].

## Data Availability

The dataset and the statistical codes supporting the conclusions of this article are openly available in the CAMHI Open Science Framework repository [http://doi.org/10.17605/OSF.IO/CRZ6H]

## Acknowledgments

The Stavros Niarchos Foundation (SNF) under its Global Health Initiative has partnered with the Child Mind Institute and a countrywide Network of mental health providers in the public system to jointly design, launch, and deliver the Child and Adolescent Mental Health Initiative (CAMHI) in Greece, which supports this work. The authors would like to thank SNF’s Co-President Andreas C. Dracopoulos and CMI’s President Dr. Harold Koplewicz for their leadership in creating, launching, and supporting this project. We would also like to thank Ms. Elianna Konialis, Ms. Dimitra Moustaka and Mr. Panos Papoulias for their critical role in multiple steps of the conceptualization and implementation of the CAMHI objectives. Finally, thanks to CAMHI hub leaders and hub members that participated in multiple stages of this work and participants and their families for their contribution.

## Ethics Declaration

### Ethical approval

Ethical approval was granted by the Research Ethics Committee of the Democritus University of Thrace [approval number: ΔΠΘ/EHΔE/42772/307].

### Consent to participate

Informed consent was obtained from all individual participants included in the study.

