## Supplemental Material for "Experience of Service Questionnaire (ESQ) in children and adolescents: factor structure, reliability, validity, item parameters and interpretability of the parent version for practical use in Greece"

### Supplementary Material

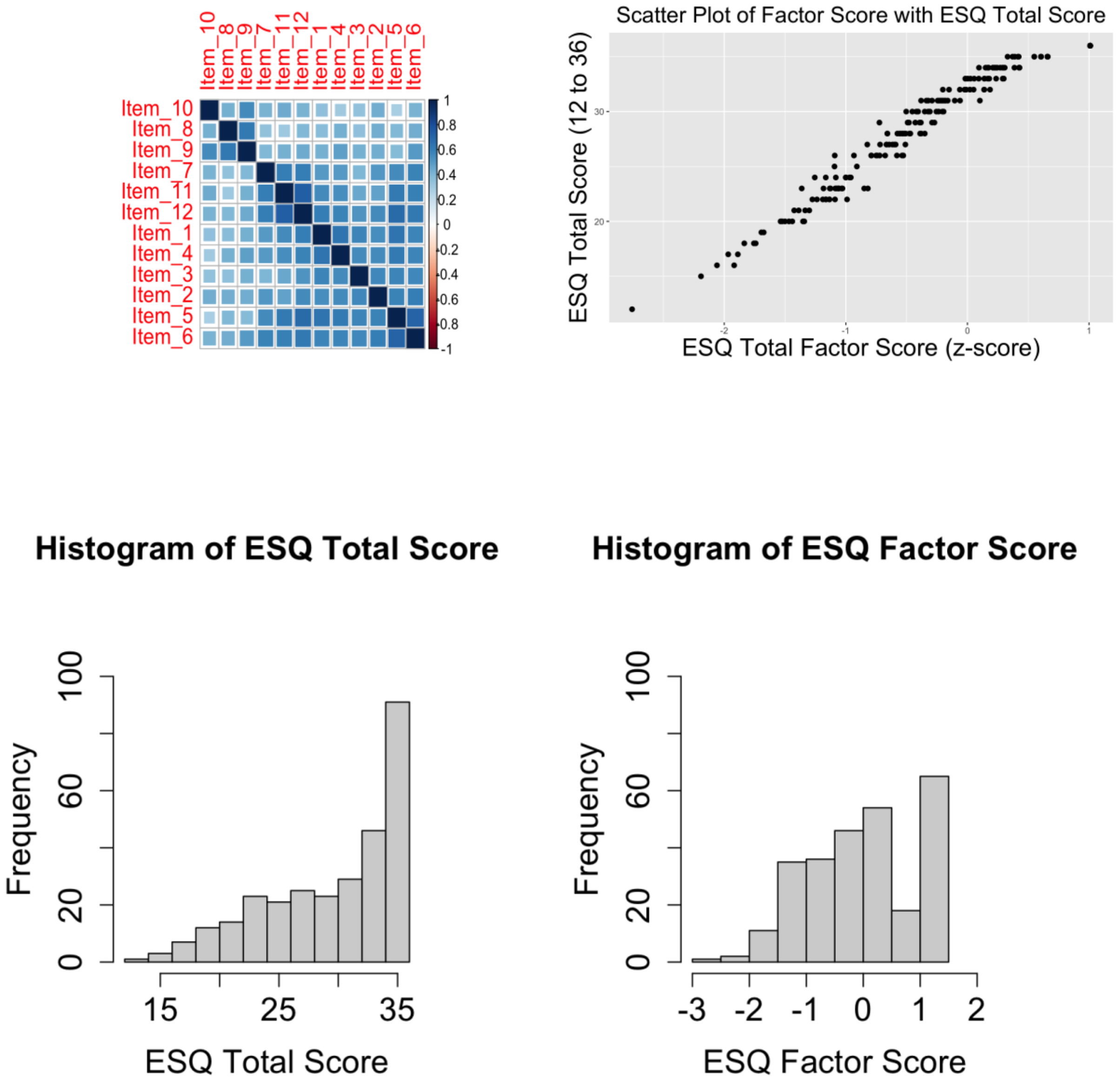


**Supplemental Figure S1.** Correlation matrix, histograms of the summed-based score and IRT-based score and scatter plot showing the association between summed score and IRT-based score

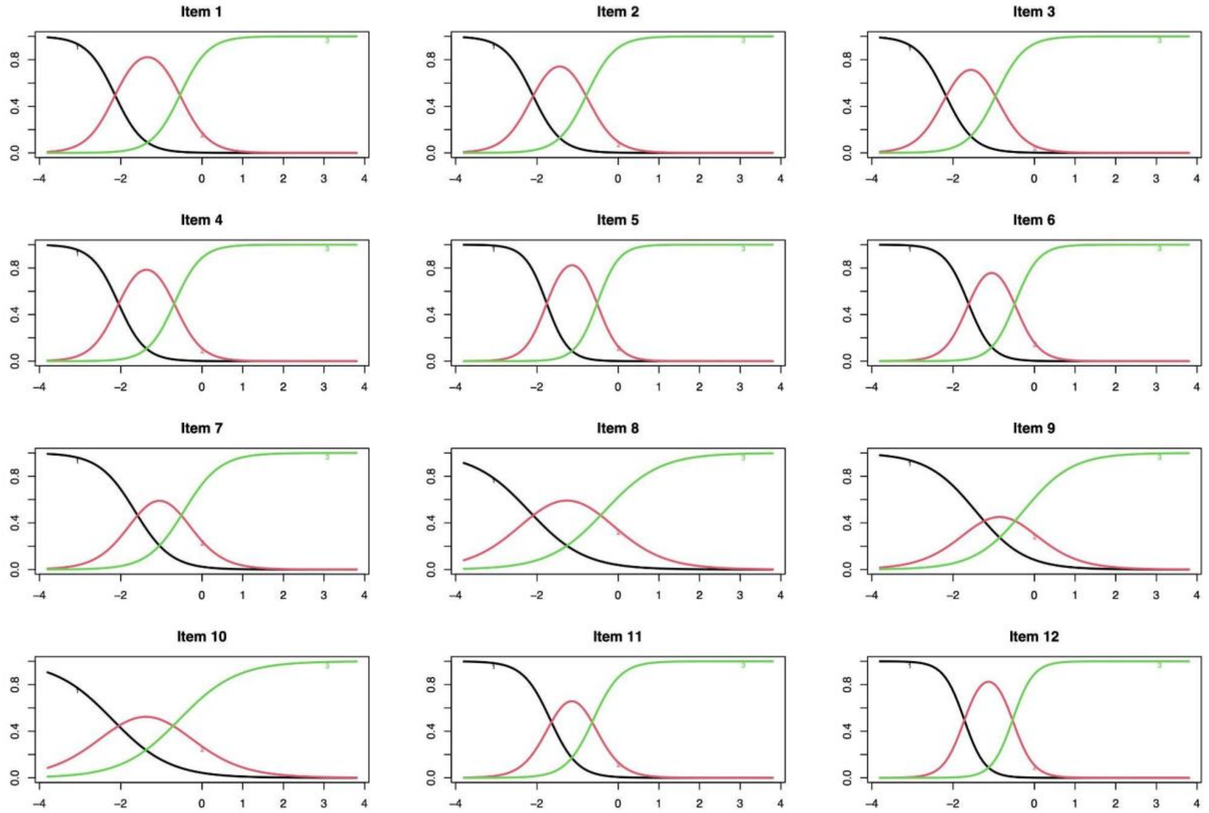


**Supplemental Figure S2:** Item Response Characteristic Curves (unidimensional solution)

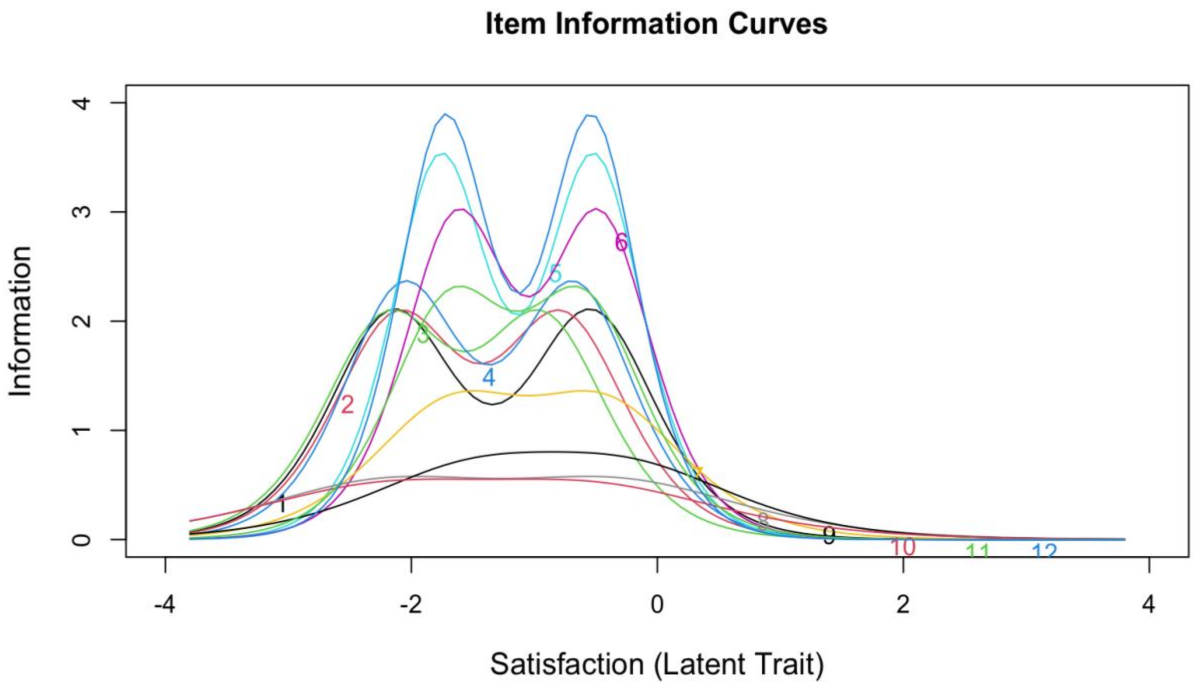


**Supplemental Figure S3.** Item Information Curves (unidimensional solution)
